# Genetic Architecture of Placental Efficiency for Term Infants: Monoaminergic Pathways and Placental Tissue Expression

**DOI:** 10.1101/2025.10.08.25337553

**Authors:** Jonas Østerhaug Andersen, Stener Nerland, Piotr Pawel Jaholkowski, Gianluca Ursini, Srdjan Djurovic, Anne Cathrine Staff, Anders Dale, Ole A. Andreassen, Ingrid Agartz, Alexey Shadrin, Laura A. Wortinger

**Affiliations:** Department of Adult Psychiatry, Division for Mental Health and Substance Abuse, Diakonhjemmet Hospital, Oslo, Norway; Division of Mental Health and Addiction, Institute of Clinical Medicine, University of Oslo, Oslo, Norway; Centre for Precision Psychiatry, Division of Mental Health and Addiction, Institute of Clinical Medicine, University of Oslo, Oslo, Norway; Lieber Institute for Brain Development, Johns Hopkins University Medical Campus, Baltimore, MD, USA; Department of Psychiatry and Behavioral Sciences, Johns Hopkins University School of Medicine, Baltimore, MD, USA; Department of Medical Genetics, Oslo University Hospital and University of Oslo, Oslo, Norway; Institute of Clinical Medicine, Faculty of Medicine, University of Oslo, Oslo, Norway; Division of Obstetrics and Gynaecology, Oslo University Hospital, Oslo, Norway; Center for Multimodal Imaging and Genetics, University of California, San Diego School of Medicine, La Jolla, CA, USA; Department of Radiology, University of California, San Diego School of Medicine, La Jolla, CA, USA; Department of Cognitive Science, University of California, San Diego, La Jolla, CA, USA; Department of Neurosciences, University of California, San Diego School of Medicine, La Jolla, CA, USA; Department of Psychiatry, University of California, San Diego School of Medicine, La Jolla, CA, USA; KG Jebsen Centre for Neurodevelopmental disorders, University of Oslo, Oslo, Norway; Centre for Psychiatric Research, Department of Clinical Neuroscience, Karolinska Institutet & Stockholm Health Care Sciences, Stockholm Region, Stockholm, Sweden; Department of Psychology, Oslo New University College, Oslo, Norway

**Author notes:** Corresponding authors: Jonas Østerhaug Andersen, Laura A. Wortinger.

## Abstract

The placenta plays a central role in supporting fetal growth. The birthweight-to-placental weight ratio is a key index of placental efficiency (PLE) and its capacity to adapt to fetal demands. We conducted the first genome-wide association study (GWAS) of PLE in 63,875 term singleton births from the Norwegian Mother, Father and Child cohort (MoBa), with complementary maternal (N = 60,472) and paternal (N = 40,116) analyses. Across offspring and maternal genomes, we identified multiple genome-wide significant loci, with TSNAX-DISC1 consistently implicated. Comparative genetic analyses showed overlap between PLE and placental weight, but minimal overlap with birth weight, indicating that PLE captures aspects of placental adaptation beyond overall growth. Gene-set enrichment highlighted monoaminergic pathways, particularly norepinephrine uptake and transport, and gene-property analyses demonstrated enrichment in placental tissue, unlike birth weight or placental weight. Global and local genetic correlation analyses revealed predominantly strong negative correlations between PLE and offspring, maternal, and paternal placental weight, strongest for paternal genomes. Notably, mapped genes including SLC6A2, SLC22A2, and SLC22A3 connect PLE to monoamine signaling, consistent with a placental contribution to neurodevelopment. These findings reveal a distinct genetic architecture of PLE and shared pathways linking placental function and offspring brain development.

## Introduction

The human placenta is a transient organ that forms the critical interface between mother and fetus. Beyond mediating the exchange of oxygen, nutrients, and waste, it supports pregnancy-specific maternal physiological processes and fetal growth (1-4). As a highly dynamic structure, the placenta adapts its morphology and nutrient transport capacity in response to both normal and adverse intrauterine environments, thereby influencing fetal development and shaping long-term health outcomes (5). Placental dysfunction has been associated with obstetric complications, including fetal growth restriction (6), preeclampsia (7, 8), and spontaneous preterm birth (9, 10), as well as adverse outcomes for offspring, including altered neurodevelopment (11, 12) and later somatic health risks (2).

Placental efficiency (PLE) can be measured as the ratio of birth weight (BW) to placenta weight (PW) (13, 14). The BW:PW ratio acts as a proxy for placental structural adaptation in response to fetal development demands (4, 13). Abnormal PLE has been associated with increased risks of adverse outcomes, with low PLE linked to fetal growth restriction (15), and childhood neurodevelopmental delay (16), and high PLE with stillbirth (17, 18). This suggests the BW:PW ratio is not merely descriptive, but also a meaningful indicator of placental adaptation to intrauterine challenges (13). As such, PLE offers valuable insights into fetal health and its developmental consequences (5).

While the genetic architecture of both PW (19) and BW (20, 21) has been studied previously, no genome wide association study (GWAS) has been performed on PLE as measured with the BW:PW ratio. Consequently, little is known about the genetic architecture of PLE itself, and whether it converges or diverges from the biology underlying placental and fetal growth. To investigate the biological basis of placental efficiency, we conducted a GWAS of the BW:PW ratio in term, singleton pregnancies from the Norwegian Mother, Father and Child cohort (MoBa). Because the placenta is primarily of fetal origin (1-3) and carries genetic contributions from both parents, analyses were performed on offspring, maternal, and paternal genotype data, with secondary sex-stratified analyses in offspring. For comparison, we also performed independent GWAS of PW and BW. These analyses enabled us to assess the genetic architecture of placental efficiency, quantify its overlap with related growth traits, and identify loci and pathways contributing to fetal development.

## Results

We analyzed data from the MoBa study (22). The offspring GWAS included 63,875 individuals (N_male_ = 32,705, N_female_ = 31,170), all born at term (38–43 weeks gestational age) from singleton pregnancies. Mean gestational age was 40 weeks (SD = 1.1). PWs ranged from 200g to 1500g (median = 675g), and BWs from 1340g to 6300g (median = 3680g). Mothers (N = 60,472) and fathers (N = 40,116) had a mean age of 29.9 (SD = 4.6) and 32.4 (SD = 5.3) years at birth respectively. PLE was defined as the ratio of BW to PW, both measured in grams. For comparison, GWAS were also performed on PW and BW in the offspring. Cohort characteristics are summarized in **Supplementary Table 1.1**.

### Genome-wide associations

All GWAS analyses were performed using REGENIE (v4.1) (23) with inverse-rank normalized phenotypes, adjusting for sex, offspring gestational age, and the first 10 genetic ancestry principal components. Sex was omitted as a covariate in the sex-stratified offspring GWAS and in the maternal and paternal GWAS. Across offspring, maternal, and paternal samples, we identified multiple genome-wide significant loci associated with PLE. Gene mapping was performed through positional mapping and gene significance was assessed through Benjamini-Hochberg false discovery rate (FDR) correction of p-values acquired through MAGMA (v1.08) (24) gene analysis.

#### Offspring GWAS of PLE

In the offspring GWAS of PLE (Manhattan plot of sex-combined offspring PLE illustrated in **Figure 1** and QQ plot shown in **Supplementary Figure 1A**) we identified 16 associated genomic loci (p = 5 × 10^-8^), comprising 20 lead and 46 independent significant single nucleotide polymorphisms (SNPs). Based on positional and functional mapping, 26 genes were mapped to the identified loci, of which 19 were significant after FDR correction (p < .05).

**Figure 1.**
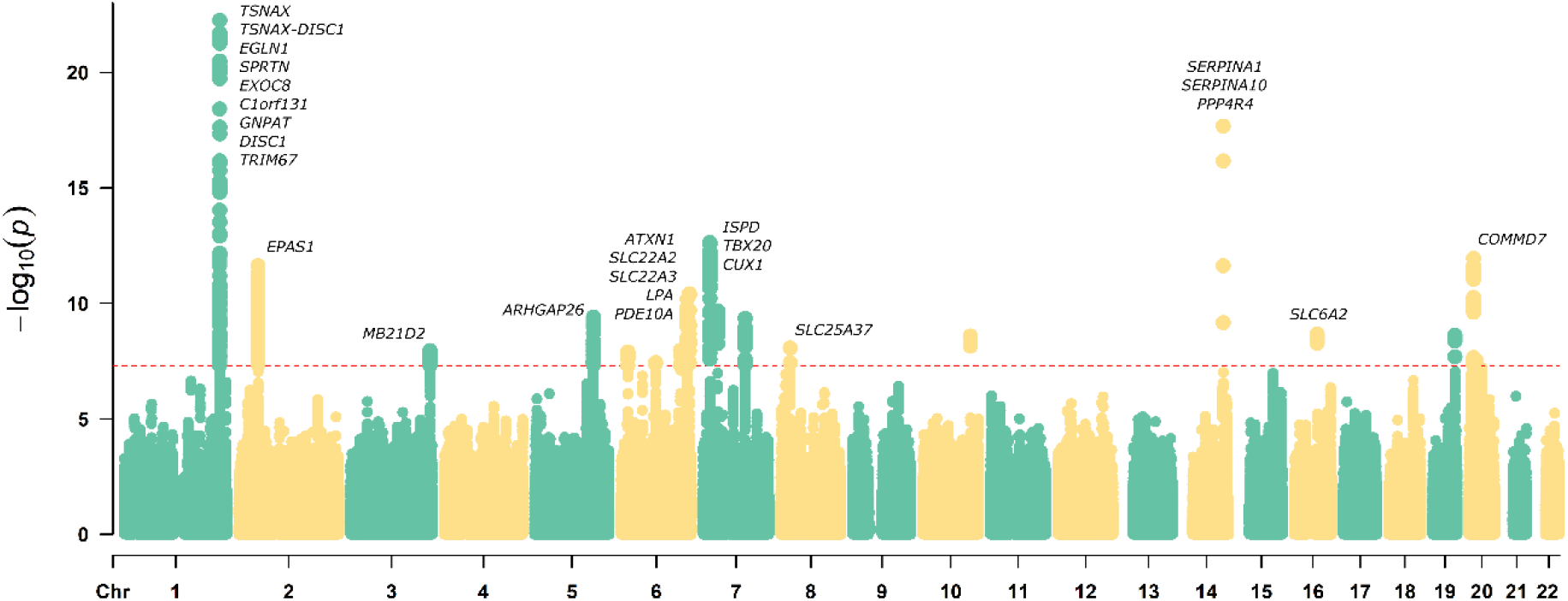
Manhattan plot with annotated genes and the significant SNPs. Each dot represents a genetic variant, plotted by chromosomal position (x-axis) and −log_10_(p) (y-axis), with higher values indicating greater statistical significance. The horizontal dashed red line denotes the genome-wide significance threshold (p = 5×10^-8^). Genes mapped from independent significant variants within associated loci are labelled.

To evaluate differences between female and male offspring, we performed a sex-stratified GWAS (Manhattan plots reported in **Supplementary Figure 2**). In females, 5 loci were detected, represented by 6 lead and 12 independent significant SNPs mapping to 8 genes of which 6 survived FDR correction (p < .05). In males, 3 loci were identified, with 4 lead SNPs, 6 independent significant SNPs, corresponding to 6 mapped genes of which 2 survived FDR correction (p < .05).

#### Maternal and Paternal GWAS of PLE

The maternal GWAS of PLE identified 4 loci (p= 5 × 10^-8^), including 4 lead and 5 independent significant SNPs mapping to 7 genes of which 3 survived FDR correction (p < .05). No significant loci were identified in the paternal GWAS (Manhattan plots reported in **Supplementary Figure 3**).

#### Placental Weight and Birth Weight GWAS of PW and BW

We performed GWAS of placental weight (PW) and birth weight (BW) in the sex-combined offspring sample using the same analysis setup, to facilitate comparison with PLE and to assess whether the associations with PLE were driven by PW or BW. In the GWAS of PW, we identified 37 genomic loci, represented by 44 lead and 100 independent significant SNPs. Gene mapping implicated 68 genes of which 19 remained significant after FDR correction (p < .05). For BW, the analysis revealed 43 loci with 50 lead and 110 independent significant SNPs leading to the mapping of 137 genes of which 96 remained significant after FDR correction (p < .05). The mapped genes identified for PW (e.g., *TSNAX– DISC1, EPAS1, ARHGAP26*) and BW (e.g., *ADCY5, CDKAL1, HMGA2*) correspond to loci previously reported in GWAS of placental weight (19) and birth weight (20). Comparison of overlap in lead SNPs and genomic loci are reported in **Supplementary Results**.

#### Shared and Unique Gene Associations Across PLE GWAS

To integrate findings, we next examined the overlap between locus lead SNPs identified in different PLE GWAS and the overlap of significant mapped genes. Overlapping genetic variants across all GWAS are reported in **Table 1**. *TSNAX-DISC1* emerged as a significant mapped gene in all PLE GWAS. *TSNAX* was present in both the sex-combined offspring and sex-stratified offspring GWAS, while *EPAS1, EXOC8, SPRTN*, and *TBX20* were shared between the sex-combined offspring and female-specific GWAS. By contrast, the maternal GWAS yielded two mapped significant genes, *MYNN* and *ACTRT3*, only present in the maternal GWAS.

**Table 1.** Overlap of lead variants between different placenta-related GWAS.

| rsID | Nearest gene | chr | pos | EA | OA | Sig SNPs | $\beta$ | p | GWAS |
| --- | --- | --- | --- | --- | --- | --- | --- | --- | --- |
| rs6659854 | *TSNAX-DISC1:*<br>*LINC00582* | 1 | 231738991 | A | C | 7 | 0.057 | 3.72E-23 | PLE, PLE Female, PLE Male, PLE Maternal, PW |
| rs139282194 | *TSNAX:*<br>*TSNAX-DISC1* | 1 | 231673221 | G | A | 8 | 0.224 | 2.98E-18 | PLE, PLE Female, PLE Male, PW |
| rs191307523 | *RNA5SP80* | 1 | 231420712 | T | G | 3 | 0.099 | 1.22E-08 | PLE |
| rs6702297 | *EGLN1* | 1 | 231541579 | C | T | 3 | 0.052 | 3.65E-08 | PLE |
| rs7571218 | *EPAS1* | 2 | 46605659 | G | A | 6 | -0.042 | 1.99E-13 | PLE |
| rs4953356 | *EPAS1* | 2 | 46578924 | G | T | 6 | -0.057 | 2.28E-13 | PLE Female |
| rs2886874 | *MB21D2* | 3 | 192657265 | T | C | 1 | 0.033 | 4.39E-09 | PLE |
| rs9841667 | *RNU6-637P* | 3 | 169438409 | T | C | 2 | 0.033 | 2.74E-08 | PLE Maternal |
| rs2651060 | *MB21D2* | 3 | 192671675 | G | A | 1 | 0.044 | 2.51E-08 | PLE Male |
| rs12187598 | *ARHGAP26* | 5 | 142402669 | C | T | 3 | 0.039 | 9.22E-10 | PLE |
| rs12209517 | *SLC22A3* | 6 | 160756736 | C | G | 3 | 0.052 | 4.28E-11 | PLE |
| rs1472265 | *PDE10A:*<br>*RP11-252P19.1* | 6 | 166182901 | T | C | 1 | -0.038 | 6.98E-11 | PLE |
| rs9367926 | *ATXN1* | 6 | 16752864 | C | G | 1 | -0.044 | 1.88E-09 | PLE, PW |
| rs10484366 | *ATXN1* | 6 | 16731311 | C | T | 1 | -0.094 | 2.07E-08 | PLE Maternal |
| rs2008133 | *ISPD* | 7 | 16211451 | C | T | 2 | 0.045 | 6.76E-15 | PLE |
| rs7790964 | *AC009531.2* | 7 | 35299884 | C | T | 1 | -0.037 | 1.70E-10 | PLE, PLE Female, PW |
| rs427534 | *CUX1* | 7 | 101886704 | A | G | 1 | 0.033 | 3.96E-09 | PLE |
| rs62503281 | *SLC25A37* | 8 | 23380743 | C | A | 1 | 0.034 | 2.81E-08 | PLE |
| rs28929474 | *SERPINA1* | 14 | 94844947 | C | T | 2 | 0.166 | 2.64E-17 | PLE, PLE Female, PLE Male, PW |
| rs11866404 | *SLC6A2* | 16 | 55717569 | C | G | 1 | -0.035 | 8.48E-10 | PLE, PW |
| rs10402234 | *MIR525* | 19 | 54199799 | C | G | 2 | 0.044 | 5.60E-12 | PLE |
| rs4499342 | *AC092566.1* | 19 | 8786913 | C | T | 1 | 0.043 | 1.50E-08 | PLE Maternal |
| rs6040470 | *RP4-734C18.1* | 20 | 11229511 | G | T | 2 | 0.041 | 2.28E-12 | PLE |
| rs6058847 | *COMMD7* | 20 | 31324135 | T | C | 1 | 0.038 | 5.51E-09 | PLE |
| rs6074298 | *RPS11P1* | 20 | 11382312 | C | T | 1 | -0.033 | 1.32E-08 | PLE |
| rs6040481 | *RP4-734C18.1* | 20 | 11237684 | G | A | 1 | 0.044 | 4.10E-08 | PLE Female |

Overlap of locus lead variants identified in the PLE GWAS from the entire offspring sample (PLE) and locus lead variants identified in maternal (PLE Maternal), male (PLE Male) and female (PLE Female), in addition to PW. GWAS were matched based on shared lead SNPs (rsID), displaying overlapping nearest genes to the SNP (Nearest gene) at each chromosome (chr) and chromosomal position (pos) in the GRCh37 genome build. BW GWAS was not included due to lack of overlap in lead SNPs. The effect allele (EA) and non-effect allele (OA) are shown for each SNP. Unique PW lead SNPs are not reported in this table. Effect sizes ( $\beta$ ), p values, and the number of independent significant SNPs in the locus (sigSNPs) for overlapping GWAS are reported from the PLE GWAS with the sex-combined offspring sample for consistency.

### Gene-set enrichment analysis of PLE

MAGMA gene-set enrichment analyses identified 9 significant pathways for the entire offspring sample and 1 significant functional pathway for male offspring, after FDR correction (**Table 2**). The top three strongest enrichments observed for the entire offspring sample were GOBP_NOREPINEPHRINE_UPTAKE (β = 1.14, p < .001), GOCC_CHROMATIN (β = 0.12, p < .05), and GOCC_CHROMOSOME (β = 0.10, p < .05) functional gene sets. The GOBP_NOREPINEPHRINE_UPTAKE set was also the single significant gene set for the male offspring sample (β = 1.7, p < .05). Of the 9 pathways identified in the sex-combined offspring sample, 5 were related to norepinephrine or other monoaminergic functions. Within these, the genes *SLC6A2, SLC22A2*, and *SLC22A3* appeared in three functional pathways and were also significant in our MAGMA gene-level analysis. No significant pathways were detected for the female offspring or maternal samples. For functional gene set analyses see **Supplementary Table 2.1-2.5**.

**Table 2.**
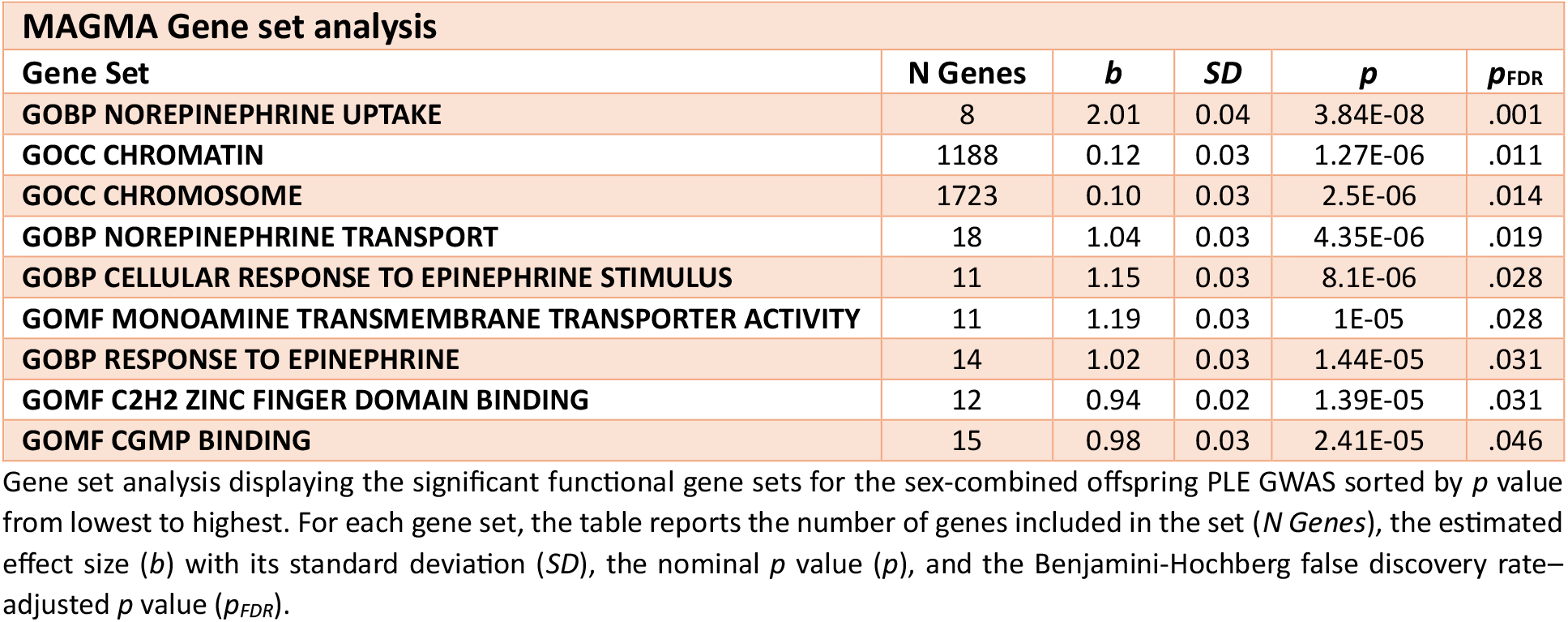
MAGMA Gene set analysis.

### Gene property analysis for tissue expression

To assess tissue-specific gene expression, we performed gene property analysis through MAGMA v1.10 (24) using all PLE GWAS as well as BW and PW GWAS, quantifying the relationship between tissue-specific gene expression and gene-phenotype association (24). Gene property analysis for tissue expression in PLE revealed significant tissue expression for the variants identified in the sex-combined offspring (**Figure 2A**), male offspring (see **Supplementary Figure 9**), and maternal sample (see **Supplementary Figure 10**). The sex-combined offspring sample exhibited significant tissue expressions in the placenta (*p* = 9.83 × 10^-5^), bladder (*p* = 1.58 × 10^-4^), esophagus gastroesophageal junction (*p* = 3.94 × 10^-4^), esophagus muscularis (*p* = 4.92 × 10^-4^), and the uterus (*p* = 5.66 × 10^-4^). The male offspring only showed a significant expression in esophagus muscularis (*p* = 8.67 × 10^-4^), while the maternal sample exhibited significant expression in both the placenta (*p* = 3.16 × 10^-4^) and the fallopian tubes (*p* = 7.08 × 10^-4^). Multiple testing was corrected using Bonferroni (p < .05). No significant gene expression was discovered for either the PW (**Figure 2B**) or BW (**Figure 2C**) GWAS.

**Figure 2.**
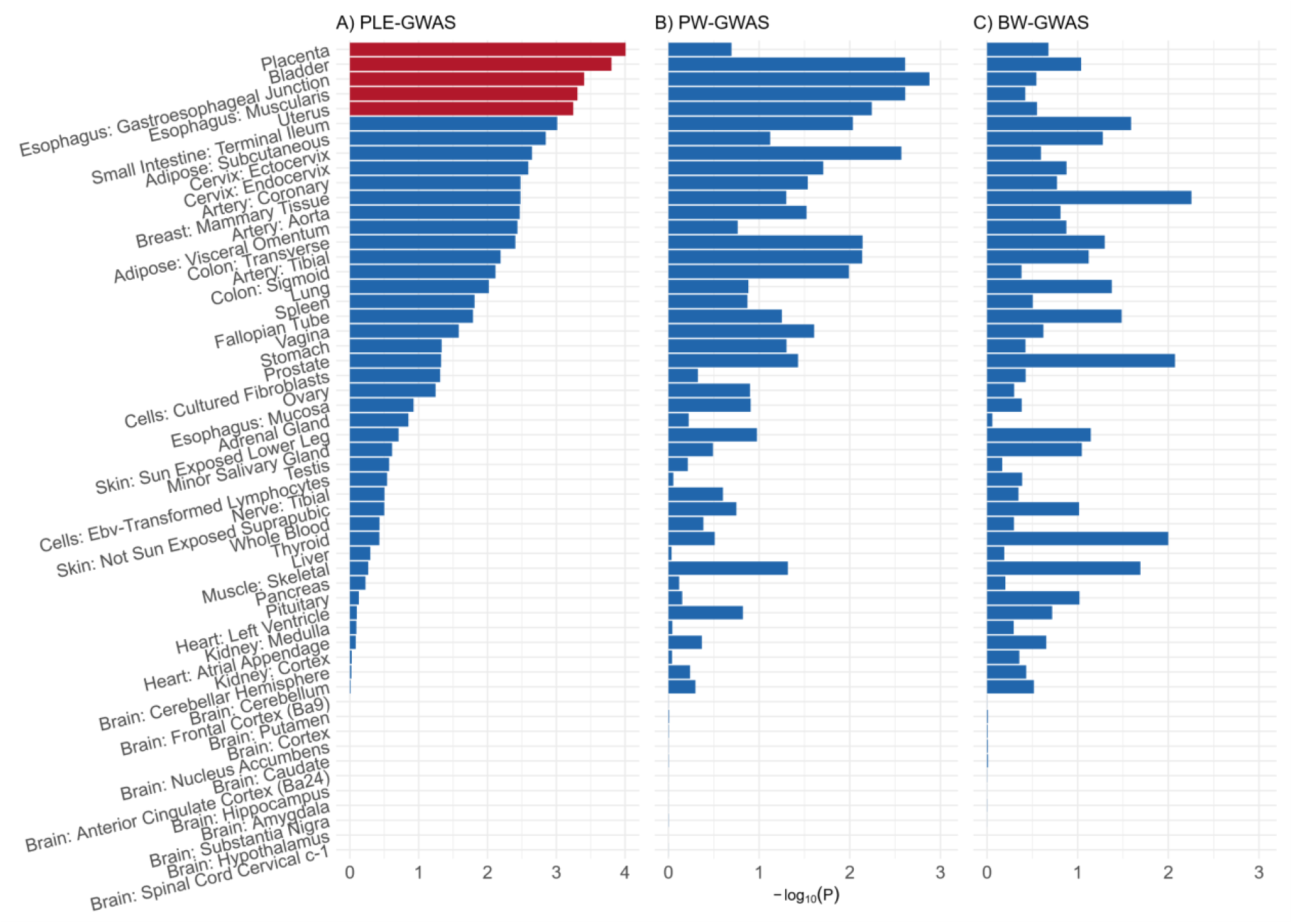
MAGMA tissue enrichment analysis. MAGMA gene property analysis was performed on (A) PLE-GWAS revealing significant expression (red bars, Bonferroni corrected p-value threshold marked with solid line) in the placenta (top), bladder, esophagus gastrointestinal junction, esophagus muscularis, and the uterus. No significant tissue enrichment was discovered in (B) PW-GWAS or (C) BW-GWAS.

### Heritability and Global Genetic Correlations

Using single-trait LD score regression (LDSC) v1.0.1 (25, 26), we estimated SNP heritability for PLE, PW, and BW. Heritability estimates were 0.109 (SE = 0.012) for PLE, 0.145 (SE = 0.014) for PW, and 0.187 (SE = 0.016) for BW.

We then used cross-trait LDSC to estimate genetic correlations between offspring PLE and GWAS traits from the Early Growth Genetics Consortium (EGG) that represent offspring, maternal, and paternal genetic components of early-life phenotypes. These included offspring and maternal BW (21), offspring, maternal, and paternal PW (19), offspring and maternal gestational age (GA) (27, 28), birth length (29), infant and birth head size, indexed by head circumference (30), and longitudinal growth measures in infancy and early childhood, indexed by the rate of change in BMI (31) (**Figure 3A**). Full results are reported in **Supplementary Table 3.1**.

**Figure 3.**
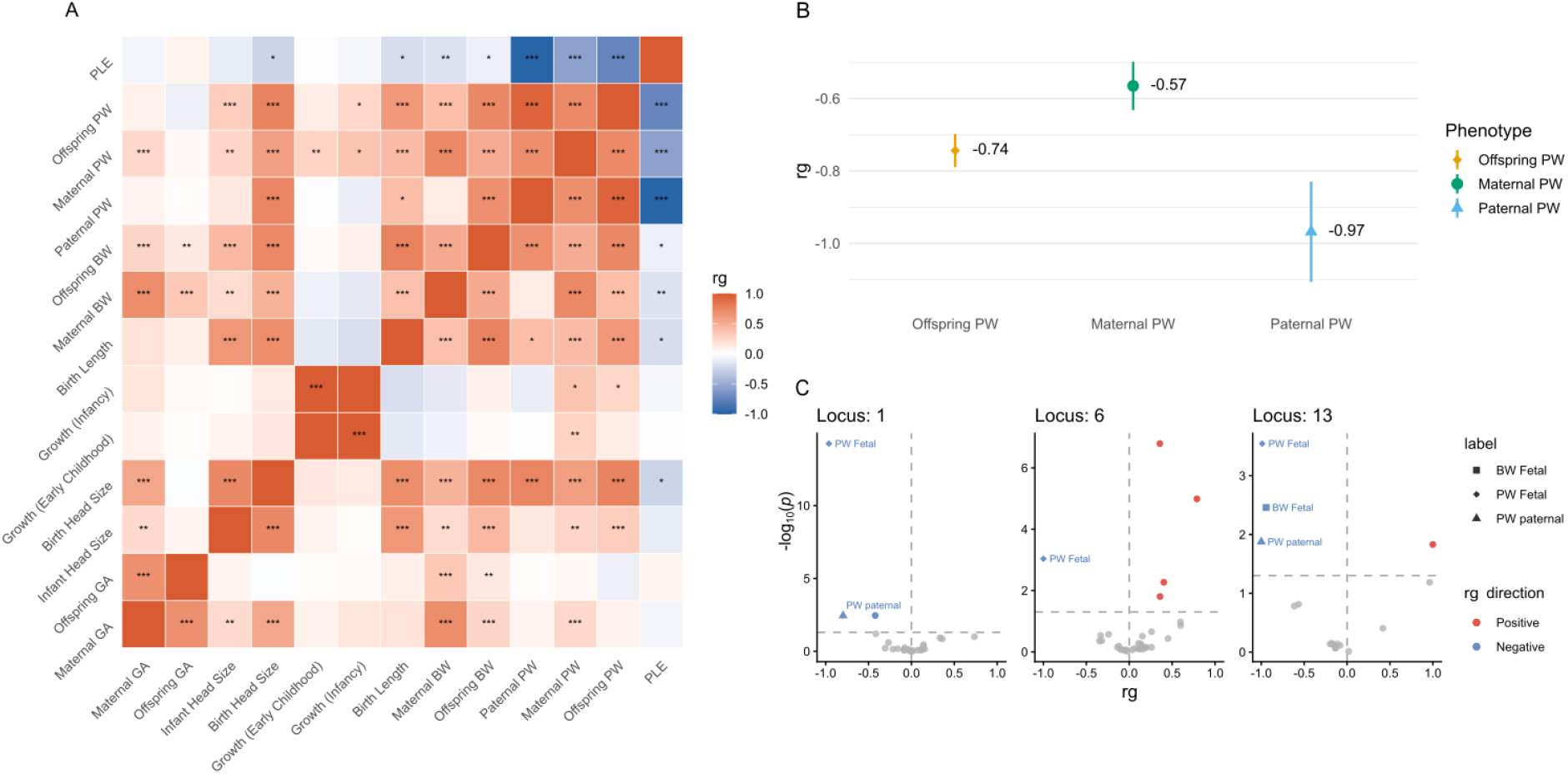
Genetic correlations between PLE and early growth phenotypes. A) Heatmap of LDSC-based genetic correlations (rg) between offspring PLE and phenotypes obtained from the Early Growth Genetics Consortium (EGG). Significant correlations are annotated within each tile (*p < .05, **p < .01, ***p < .001). B) Dot-and-whisker plots of offspring PLE genetic correlations with offspring, paternal, and maternal PW. Strong negative genetic correlations were observed for offspring (yellow), maternal (green), and paternal (blue) genetic components. C) Volcano plots of local genetic correlations in selected loci identified in the offspring PE GWAS, computed using LAVA. The x-axis shows local rg, and the y-axis shows -log10(P) values. Significant negative correlations are shown in blue and significant positive correlations are shown in red. Vertical and horizontal gray dashed lines indicate the local rg = 0 and the Benjamini-Hochberg FDR-corrected significance threshold of p < .05, respectively.

We observed strong negative correlations between offspring PLE and offspring PW (rg = –.74, SE = .05, p < .001), maternal PW (rg = –.57, SE = .06, p < .001), and paternal PW (rg = –.97, SE = .14, p < .001). Correlations varied across offspring, maternal, and paternal genetic components, potentially reflecting distinct genetic contributions to the relationship between PLE and placental growth (**Figure 3B**). More modest, but statistically significant, negative correlations were observed between offspring PLE and head size at birth (rg = –.25, SE = .11, p < .05), offspring BW (rg = –.09, SE = .04, p < .05), and maternal BW (rg = –.16, SE = .06, p < .01).

### Local Genetic Correlations

To assess whether genome-wide genetic correlations were accompanied by local genetic correlations at mapped PLE loci, we performed local genetic correlation analyses between PLE and EGG traits using Local Analysis of [co]Variant Association (LAVA v0.1.5) (32) (**Figure 3C, Supplementary Table 3.2**). Analyses were restricted to the 16 PLE-associated loci (**Supplementary Figure 4**), allowing us to evaluate whether PLE-associated regions showed evidence of local genetic correlation between offspring PLE and early growth phenotypes. Local genetic correlations were estimated only for trait pairs showing significant local heritability at the same locus. Pairwise correlations were corrected for multiple testing using the Benjamini-Hochberg procedure.

Among the 16 PLE-associated loci, all loci except locus 3 showed at least one significant local genetic correlation with PLE. Consistent with the genome-wide LDSC analyses, most significant local correlations involved offspring, maternal, and paternal PW, as well as offspring BW, and these correlations were consistently negative. In addition, we identified significant negative local genetic correlations with offspring genetic effects on head size at birth in locus 15 and infant head size at locus 8. Positive local genetic correlations were observed between PLE and offspring gestational age at locus 11 and maternal gestational age at locus 15, despite no significant global genetic correlation for these trait pairs. These findings suggest that the pattern of local genetic correlations between PLE and EGG phenotypes might consist of an intricate mixture of regions with concordant and discordant effects.

## Discussion

We present the results from GWAS of PLE based on the BW:PW ratio. Across offspring, maternal, and paternal genomes, we identified multiple loci associated with PLE in the maternal and offspring GWAS, but no significant paternal associations. Gene-based analyses revealed several significant genes, notably *TSNAX-DISC1*, which was consistently implicated across all PLE GWAS, apart from the paternal GWAS. Genetic correlation analyses revealed substantial, predominantly negative relationships between PLE and PW, across offspring, maternal, and paternal components, but only modest correlations with BW, head size, and birth length. Local genetic correlation analyses further indicated shared genetic architecture in PLE-associated loci. Functional pathway analyses further highlighted strong enrichment for norepinephrine and monoaminergic signaling, supported by convergent evidence from mapped and significant genes such as *SLC6A2, SLC22A2*, and *SLC22A3*. We also demonstrated significant placental expression of PLE genes, which was not the case for PW or BW GWAS.

One consistent finding across our PLE GWAS was the involvement of *TSNAX-DISC1*, which was mapped in the fetal, maternal, and sex-stratified analyses. *TSNAX-DISC1* is a read-through transcript formed when *TSNAX* transcription continues into *DISC1*. Importantly, in our analyses, *TSNAX* was mapped and significant both in the sex-combined offspring GWAS and in sex-stratified GWAS, while *DISC1* was significant only in the sex-combined offspring sample. The *TSNAX* gene encodes for the protein Translin-associated factor X which binds to the translin protein and has been implicated in DNA repair functions and neural plasticity (33). *DISC1* has a key role in early neurodevelopment, synaptic regulation and cognitive health (34-38) and it also been associated with several functions such as coordination of intracellular trafficking which in turn affects neuronal development and connectivity (39), dopamine regulation (40), and synaptic plasticity (38). Prior and recent (41) studies have implicated this region in psychiatric risk: the *TSNAX-DISC1* locus shows association with disorders such as schizophrenia and other psychiatric disorders and related traits (34, 42-47). Notably both *TSNAX* and *DISC1* are expressed in the placenta (48, 49) and recent work has shown that DISC1, in placenta, activates autophagy by enhancing AMPKα phosphorylation and reducing mTOR phosphorylation, protecting against infection (50), consistent with findings in neural progenitor cells (51). Furthermore, *TSNAX–DISC1* has been identified as a chimeric transcript that may promote endometrial carcinoma progression (52), consistent with similarities between trophoblast development and cancer invasion. Furthermore, our LAVA analysis of PLE-associated loci revealed strong local genetic correlations between offspring PLE and offspring and paternal PW, with weaker correlations for maternal PW. This suggests that genetic overlap between PLE and placental growth at PLE-associated loci may be more strongly aligned with offspring and paternal genetic components, with relevance for placental pathways influencing fetal development. The convergence of placental expression, neuropsychiatric association, and cancer-related dysregulation supports the hypothesis that regulation at the *TSNAX-DISC1* locus could act as a developmental link between placental inefficiency, offspring vulnerability to psychiatric disorders.

We observed strong negative genetic correlations between PW and PLE, alongside substantial overlap of significant genes and loci. This indicates that PLE and PW share part of their underlying genetic architecture. The particularly strong negative correlation with paternal PW further suggests that PLE may be especially aligned with paternal genetic components of placental growth.

Local genetic correlation analyses supported the genome-wide LDSC findings. Most PLE-associated loci exhibited significant local genetic correlations, the majority involving offspring, maternal, or paternal PW, and these were predominantly negative. In contrast, the weaker negative correlation between PLE and BW points to a more limited genetic relationship with fetal growth. Large-scale BW GWAS have shown that BW reflects both direct fetal genetic effects and indirect maternal genetic effects, including pathways related to fetal insulin secretion, maternal glucose, maternal height, and blood pressure (21). The limited genetic overlap between BW and PLE suggests that these GWASs may capture partly distinct genetic signals. However, without functional validation, we cannot determine whether the observed PLE- or BW-associated signals reflect placental function directly. This interpretation is however consistent with the stronger shared global and local genetic architecture between PLE and PW, which may reflect placental hypertrophy or compensatory placental growth (53).

Despite negligible genome-wide genetic correlations for several early-growth traits, we identified locus-specific correlations with BW, head size, birth length, and gestational age that were not always reflected in the LDSC estimates. Although offspring PW largely dominated PLE local correlations, paternal PW emerged as a major contributor. Paternal and maternal local effects overlapped at only two loci, and we observed more paternal than maternal PW correlations overall. These contrasting correlation patterns, alongside differences in the strength of their global genetic correlations with PLE, are compatible with literature implicating parent-of-origin effects in placental development, fetal growth, and fetal-maternal resource allocation. In this framework, paternally expressed genes are often associated with enhanced placental and fetal growth, whereas maternal genetic effects may also partly reflect constraints imposed by maternal physiology and the intrauterine environment (54-57). Although our results do not directly establish imprinting mechanisms, they suggest that parent-of-origin effects may be a useful framework for future analyses.

Despite global and local overlap between PW and PLE, the lead SNPs driving PLE associations were largely unique (**Supplementary Results, Supplementary Figure 5**), implying that PLE reflects distinct aspects of placental function not fully captured by PW. This distinction aligns with a recent GWAS of PW (19), which revealed heterogeneous architecture with predominant fetal contributions but additional maternal and paternal influences, spanning pathways in placental development, nutrient and antibody transport, immune function, and metabolism, and showing partial overlap with BW signals. While PW reflects diverse and partly nonspecific influences on placental mass, PLE may help refine genetic signals related to placental adaptation, making it a more targeted phenotype for studying placental biology and its downstream implications for offspring development.

The sex-combined offspring GWAS revealed nine significant functional gene sets, where one gene set was shared with the male offspring GWAS. No other GWAS revealed functional gene sets. Of these gene sets, five were associated with norepinephrine (NE) and monoamine functions. Within these functional groups three of them contained the *SLC6A2, SLC22A2*, and *SLC22A3* genes, which were mapped and significant in the sex-combined GWAS. *SLC6A2* codes for the NE transporter NET (58), which mainly transports NE through cellular membranes, but has also been shown to be permeable to dopamine and serotonin (59). This is also the case for *SLC22A2* and *SLC22A3* which codes for the OCT2 and OCT3 proteins respectively. OCT3 has been shown to regulate NE, serotonin, and dopamine (59-62), while OCT2 has been shown to mainly be permeable towards epinephrine, but exhibits an affinity towards dopamine and NE, though to a lesser extent (63, 64). Notably, *SLC6A2* has been implicated in neurodevelopmental and psychiatric phenotypes in offspring, most prominently ADHD, including gene-environment interactions involving maternal smoking during pregnancy (65-67). This highlights the broader relevance of placental monoaminergic regulation for fetal neurodevelopment. At the same time, *SLC6A2* has also been identified among genes predictive of early preeclampsia (68), linking monoamine transport to pregnancy complications characterized by vascular dysfunction and altered maternal-placental physiology. Furthermore, maternal-facing NET and fetal-facing OCT3 have been identified in the syncytiotrophoblast, where they regulate monoaminergic concentrations in maternal and fetal circulations (59). In addition, the significant local genetic correlation between offspring PLE and paternal PW at the *SLC6A2* locus suggests a potential mechanism by which paternal genetic effects on placental growth may influence monoaminergic regulation at the fetal-maternal interface. This provides a plausible route for paternal genetics to impact both fetal developmental outcomes, and maternal health during pregnancy. Together, these findings support a model in which PLE-associated monoamine transport genes contribute to placental function, offspring neurodevelopmental outcomes, and maternal health.

Gene property analysis revealed significant expression in the placenta, bladder, esophagus, and uterus in the sex-combined offspring sample. In male offspring, the gene property analysis revealed significant gene expression in the esophagus muscularis tissue, while also discovering significant gene expression in the placenta and fallopian tubes in the maternal GWAS. Placental enrichment aligns with the central role of placental biology in PLE, while uterine and fallopian tube signals in the maternal genome underscore the contribution of maternal reproductive tissues. This is consistent with previous studies implicating *TSNAX-DISC1* in endometrial carcinomas (52), *SLC6A2* and *SLC22A3* expression in the syncytiotrophoblast of the placenta (59), and with increased *SLC6A2* expression during gestation (69). The absence of significant signals in PW and BW GWAS further suggests that these expression patterns are either unique to PLE or offers a more specific genetic structure, supporting a distinct biological profile despite overlapping global architecture with related PW traits.

The developmental context of monoaminergic signaling provides a plausible mechanism linking these placental processes to offspring brain development. During early neurodevelopment, the fetal brain expresses monoamine receptors before intrinsic monoaminergic neurons mature, while endogenous monoamine production remains low throughout gestation (70). Despite the enrichment of PLE genes for monoaminergic pathways, we did not observe significant expression of PLE genes in the fetal brain or neuronal tissues. Although monoamine production increases toward term (71), the placenta is likely the sole source of serotonin during gestation (72, 73), while native fetal catecholamine concentrations remain limited (70). The developing fetus is therefore dependent on placental monoamine synthesis and regulation, as fetal monoamine availability is largely shaped by placental processing of maternal monoamines (69, 72-74). Furthermore, catecholaminergic transporters are mainly expressed in the placenta, whereas their expression in fetal tissues is generally low (59, 69). Expression of the NET transporter in the human placenta increases from the first trimester toward term, while no corresponding developmental change is observed in the fetal brain (69). The low capacity for fetal monoamine production, and the limited expression of monoaminergic transporters in fetal tissues, may explain the absence of detectable PLE gene expression in cerebral tissue, in contrast to their robust expression in the placenta.

Our PLE GWAS findings suggest important developmental and implications by associating placental regulation of monoaminergic pathways in offspring neurodevelopment. The strong genome-wide negative genetic correlations between PLE and PW, together with modest or negligible correlations with BW and other early growth traits, indicate that PLE is more closely related to genetic variation in placental growth and adaptation rather than to generalized fetal growth. The enrichment of NE related gene sets, alongside signals in genes such as *TSNAX-DISC1*, points to a potential mechanism by which placental adaptation influence vulnerability to neuropsychiatric disorders. Shared genetic signal between PLE and PW was evident at multiple PLE-associated loci, and local correlations were often strongly negative, particularly for offspring and paternal PW. This suggests that the genetic relationship between placental growth and efficiency is shaped by specific genomic regions rather than by uniform genome-wide effects. In this context, the paternal PW correlations are particularly relevant. LAVA also identified limited overlap between maternal and paternal local effects, supporting the idea that maternal and paternal genetic contributions may influence placental adaptation through partly distinct biological pathways. This aligns with emerging literature linking placental genetic risk to schizophrenia (75-77), studies linking schizophrenia risk to altered DNA methylation (78) and placental gene expression (78, 79), as well as reports of neurodevelopmental alterations associated with abnormal PLE ratios (16). The overlap between paternal PW correlations and monoaminergic loci also suggests a possible link between paternal genetic components and placental pathways involved in monoaminergic signalling, with potential relevance for offspring brain development. Expression studies further support this interpretation as NET and OCT3 expression are significantly reduced in pre-eclamptic placentae, suggesting that impaired monoamine clearance may contribute to elevated circulating norepinephrine, placental vasoconstriction, and reduced fetal perfusion (80). Furthermore, monoamine transporters from multiple families have been identified in endometrium, decidua, and placenta, where they regulate extracellular monoamine concentrations essential for implantation, placentation, and fetal development (81).

Given the placenta’s role in synthesizing and regulating monoamines (59, 69, 74), our results suggest a dual function whereby the placenta supports its own health (80, 81) while also influencing neuronal migration, proliferation, and differentiation (70, 72), highlighting a direct genetic connection between placental monoamine function and offspring neural development. Together, these findings suggest common genetic pathways may contribute to both placental function and neurodevelopmental processes in the offspring (12, 74, 80, 81). Our results provide new insight into potential shared biological basis between placental adaptation and neurodevelopment, consistent with the placenta’s role as a key interface between maternal and offspring biology.

## Materials and Methods

### Participants and phenotype definition

The sample is a subset of the Norwegian Mother, Father and Child cohort study (MoBa; https://www.fhi.no/en/ch/studies/moba/). MoBa is a population-based pregnancy cohort study conducted by the Norwegian Institute of Public Health of mothers, fathers, and their children. Participants were recruited across Norway from 1999-2008. The women consented to participation in 41% of the pregnancies. The cohort includes approximately 114,500 children, 95,200 mothers and 75,200 fathers. MoBa is regulated by the Norwegian Health Registry Act. The current study was approved by The Regional Committees for Medical and Health Research Ethics (2016/1226). Pregnancy-related factors were acquired through the Medical Birth Registry of Norway (MBRN). The MBRN is a national health registry containing information about all births in Norway. We analyzed data from single birth, at term births (38 weeks – 43 weeks gestational age), and placental weights ranging from 200g-1500g. Pregnancies where the fetus had genetic syndromes or structural acknowledged anatomical malformations were excluded. GWAS were performed on offspring (N = 63,894, N_male_ = 32,711, N_female_ = 31,183), maternal (N = 60,472), and paternal (N = 40,116) samples, all white-European genetic ancestry. Placental efficiency was computed as the birth weight-placenta weight (BW:PW) ratio. GWAS was also performed on birth weight and placental weight for the fetal sample, for the purpose of genetic correlation analysis.

### Genome-wide association analysis

#### Genotyping and QC

Blood samples were collected from mothers and fathers around the 17th week of gestation, and additional samples were obtained from the mother postpartum and from the child’s umbilical cord at birth. DNA was extracted at the Norwegian Institute of Public Health using standard procedures (https://www.fhi.no/en/publ/2012/protocols-for-moba/). Genotyping was conducted across several research projects over multiple years, resulting in 238,001 samples processed across 26 technical batches using different arrays and genotyping centers. Some individuals were genotyped more than once for quality control or due to overlapping projects (for details see (82)).

Genotype quality control and imputation were conducted using the standardized MoBaPsychGen pipeline v.1(62). The MoBaPsychGen pipeline is a 9-module pipeline, accounting for complex relatedness structures. Variants with low imputation quality were excluded resulting in 6,981,748 high-quality common autosomal variants from 207,569 unique individuals (82).

#### GWAS

Genome-wide association analyses were performed using REGENIE (v4.1; (23)), a two-step whole-genome regression method. In Step 1, we used ∼500,000 variants that were either genotyped or imputed with very high quality. Variants were filtered for minor allele frequency (MAF) ≥ 1%, genotype missingness ≤ 10%, and Hardy–Weinberg equilibrium p ≥ 1 × 10^−15^. This step was used to fit the prediction model for phenotypes while accounting for relatedness and population structure. In Step 2, we tested all imputed variants passing the following filters: minor allele count (MAC) ≥ 20 and imputation INFO score ≥ 0.8. Association analyses were conducted using linear regression adjusted for sex, gestational age, and the first 10 genetic ancestry principal components. All phenotypes were inverse-rank normalized prior to analysis.

#### Genomic Risk Loci and Lead SNPs

Genomic risk loci were identified using the SNP2GENE function in Functional Mapping and Annotation of Genome-Wide Association Studies (FUMA) v1.5.2 (83). Independent significant SNPs were defined as those with a P-value < 5 × 10^−8^ and in low linkage disequilibrium (r^2^ < 0.6) with one another. Among these, lead SNPs were identified as a subset with r^2^ < 0.1. Genomic risk loci located within 250 kilobases of each other were merged and considered a single locus. 1000 Genomes Phase 3 European reference panel panel was used to estimate linkage disequilibrium.

#### Functional mapping and annotation

Functional mapping and annotation were conducted using FUMA. Independent significant SNPs (p < 5 × 10^−8^) and their LD-expanded candidate SNPs (r^2^ ≥ 0.6, based on the 1000 Genomes Phase 3 European reference panel, EUR population) were annotated using ANNOVAR (2017-07-17) with Ensembl build v102 gene definitions. Functional scores were assigned to candidate SNPs using Combined Annotation Dependent Depletion (CADD), RegulomeDB (RDB), and 15-core chromatin state annotations from the Roadmap Epigenomics Project. SNPs were mapped to protein-coding genes using positional mapping, defined as ±10 kb from the transcription start site of each gene.

### Genome Wide genetic correlation analysis using LDSC

To assess genetic similarity between PLE and EGG-derived phenotypes we performed genetic correlation analysis using linkage disequilibrium score regression (LDSC) (25, 26). Birth Length statistics from GRCh36 was lifted over to GRCh37 genome build using the 1000 Genomes Phase 3 reference panel, by matching variants based on rsID and updating chromosome and base-pair positions to their corresponding GRCh37 coordinates. Variants were processed chromosome-wise in parallel and restricted to biallelic SNPs, after which alleles were aligned to the reference panel while accounting for for reference-alternative allele swaps and strand flips. Variants with unresolved allele mismatches were excluded. Head circumference GWAS had missing rsIDs, which were reimputed using chromosome, position, and allele information from the same GRCh37 reference panel, allowing matching in both forward and reversed allele orientations. This procedure resulted in successful rsID recovery for approximately 96.6% of SNPs while ensuring that all retained variants were consistently aligned to the GRCh37 reference build.

### Local genetic correlation analysis using LAVA

Local genetic correlation analysis was performed using LAVA (Local Analysis of [co]Variant Association, v0.1.0) (32), which tests for genetic correlations within specific genomic regions rather than globally across the genome. This approach allows for the detection of locus-specific genetic relationships that may be obscured or absent at the genome-wide level. We tested PLE-associated loci using genomic coordinates defined by FUMA (see Genomic Risk Loci and Lead SNPs section above). Sampling correlations to account for potential sample overlap between GWAS were estimated using the cross-trait LDSC intercept from cross-trait LD score regression, following the approach recommended by the LAVA developers. The European 1000 Genomes Phase 3 reference panel (g1000_eur) was used for all analyses. For each locus, a univariate test was first performed to assess whether there was sufficient local heritability for each phenotype. Bivariate local genetic correlations were subsequently estimated between PLE and all phenotypes obtained from the EGG consortium at loci passing the univariate significance threshold. Multiple testing was corrected using the Benjamini-Hochberg procedure.

### Top hit SNP and loci comparison

To further quantify genetic similarity between PLE, BW, and PW, we compared the genetic structure by assessing similarity in lead SNPs. To evaluate overlapping lead SNPs and genomic risk loci we computed Jaccard index

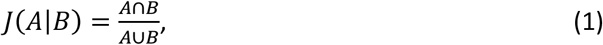

and the Szymkiewicz–Simpson overlap coefficient

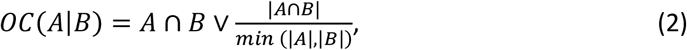

For this purpose, each genomic risk locus was defined by its base-pair span (start to end coordinates). The indices were calculated based on the total number of overlapping base pairs across loci.

To further assess overlap between loci across GWASs, we represented each locus by its start and end coordinates as defined by FUMA. For each PLE locus, we examined whether at least one genome-wide significant SNP (p < 5×10^−8^) from the PW GWAS fell within the locus boundaries. Loci containing ≥1 significant SNP from the second GWAS were counted as shared; otherwise, they were considered unique to PLE. Since all GWASs were performed in MoBa using nearly identical SNP sets, locus boundaries were approximated using the set of candidate SNPs reported by FUMA. Each locus contributed equally (*w* = 1) to the final proportion of shared versus unique loci.

Genetic overlap was also assessed descriptively and visualized through Venn diagrams, where we visually inspected the intersection of FDR significant genes between PLE and PW. FDR was performed on genes based on raw p-values obtained through MAGMA gene analysis (described below). All results from SNP and loci comparison are reported in **Supplementary Results and Supplementary Figure 5**.

### MAGMA analysis

Using Multi-marker Analysis of GenoMic Annotation (MAGMA) (24) we performed gene analysis, gene-set analysis and gene-property analysis for tissue specificity, where both gene analysis and gene-set analysis were performed using FUMA v1.5.2 using the MAGMA v1.08 implementation. Gene-property analysis for tissue specificity was performed locally using MAGMA (v1.10).

#### MAGMA gene analysis

GWAS summary statistics were input into MAGMA with SNP-level p-values. SNPs were mapped to protein-coding genes based on their physical location with no additional upstream or downstream window. Linkage disequilibrium (LD) between SNPs was accounted for using the 1000 Genomes Phase 3 European reference panel. Gene-level p-values were calculated by aggregating SNP-level summary statistics within each gene while correcting for LD structure.

#### Gene enrichment analysis

Gene enrichment analysis was conducted using MAGMA, including both functional gene set and tissue-specific expression analyses. Functional gene set analysis was conducted through the FUMA implementation of MAGMA which tests 17023 predefined biological pathways (MsigDB v2023.1). Post hoc Benjamini-Hochberg FDR correction was subsequently performed in R.

#### Gene property analysis for tissue specificity

Gene property analysis for tissue specificity was not performed through FUMA, as FUMA’s implementation of MAGMA is based on GTEx data, where placental tissue was not available by default. To include placenta, we obtained gene expression data from the POPS study through the POPS Placenta Transcriptome project (https://www.obgyn.cam.ac.uk/placentome/), described in Gong et al. (2021). The transcriptomic data was harmonized through converting Fragments per kilobase million (FPKM) to transcripts per million (TPM; (84))

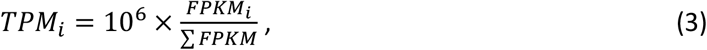

subsequently winsorized,

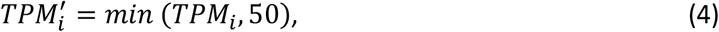

and log transformed

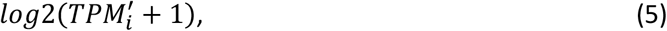

as reported in FUMA preprocessing prior to MAGMA tissue enrichment analysis. Missing gene expression values for placental tissue were replaced with the minimum expression observed for the corresponding gene across all tissues. This conservative imputation assumes that missing values reflect low gene expression and minimizes inflation of tissue-specific effects. Tissue enrichment analysis was then performed using MAGMA, using GTEx v8 with 54 and 30 tissues, alongside the placental tissue.

## Supporting information

Supplemental Results

## Data Availability

The data that support the findings of this study are derived from the Norwegian Mother, Father and Child Cohort Study and the Medical Birth Registry of Norway, both managed by the Norwegian Institute of Public Health (NIPH). Individual-level data are protected under the Norwegian Health Registry Act and are therefore not publicly available. Access to data requires approval from the Regional Committees for Medical and Health Research Ethics and from the data owners. Qualified researchers may apply for data access through the Health Research Portal of Norway.

## Acknowledgements

We are grateful to all the participating families in Norway who take part in this on-going cohort study. For generating high-quality genomic data, we thank the Norwegian Institute of Public Health (NIPH), the HARVEST collaboration, the NORMENT Centre at the University of Oslo, the Center for Diabetes Research at the University of Bergen, deCODE Genetics, the Research Council of Norway, the South-Eastern and Western Norway Regional Health Authorities, the ERC AdG, Stiftelsen KG Jebsen, the Trond Mohn Foundation, and the Novo Nordisk Foundation. We thank the Medical Birth Registry of Norway for providing data, and the participants whose data made this research possible. This work was performed on the TSD (Services for Sensitive Data) facilities, owned by the University of Oslo, operated, and developed by the TSD service group at the University of Oslo, IT-Department (USIT). Computations were also performed on resources provided by UNINETT Sigma2—the National Infrastructure for High Performance Computing and Data Storage in Norway. We would also like to thank Akira Sawa for helpful discussions regarding the DISC1 literature.

## Code Availability

Code used in this study is available via the following resources: FUMA (https://fuma.ctglab.nl/), LAVA (https://github.com/josefin-werme/LAVA), LDSC (https://github.com/bulik/ldsc), MAGMA (https://cncr.nl/research/magma/), Regenie (https://rgcgithub.github.io/regenie).

## Author contributions

J.Ø.A.: Formal analysis, Methodology, Visualization, Writing - original draft, Writing - review & editing. S.N.: Conceptualization, Writing - review & editing. P.P.J.: Data curation, Resources, Writing - review & editing. G.U.: Resources, Writing - review & editing. S.D.: Resources, Supervision, Writing - review & editing. A.C.S.: Resources, Writing - review & editing. A.D.: Writing - review & editing. O.A.A.: Funding acquisition, Resources, Writing - review & editing. I.A.: Conceptualization, Supervision, Writing - review & editing. A.S.: Methodology, Formal analysis, Writing - review & editing. L.A.W.: Conceptualization, Funding acquisition, Methodology, Supervision, Writing - review & editing.

