## Supplementary figures for "Genetic Architecture of Placental Efficiency for Term Infants: Monoaminergic Pathways and Placental Tissue Expression"

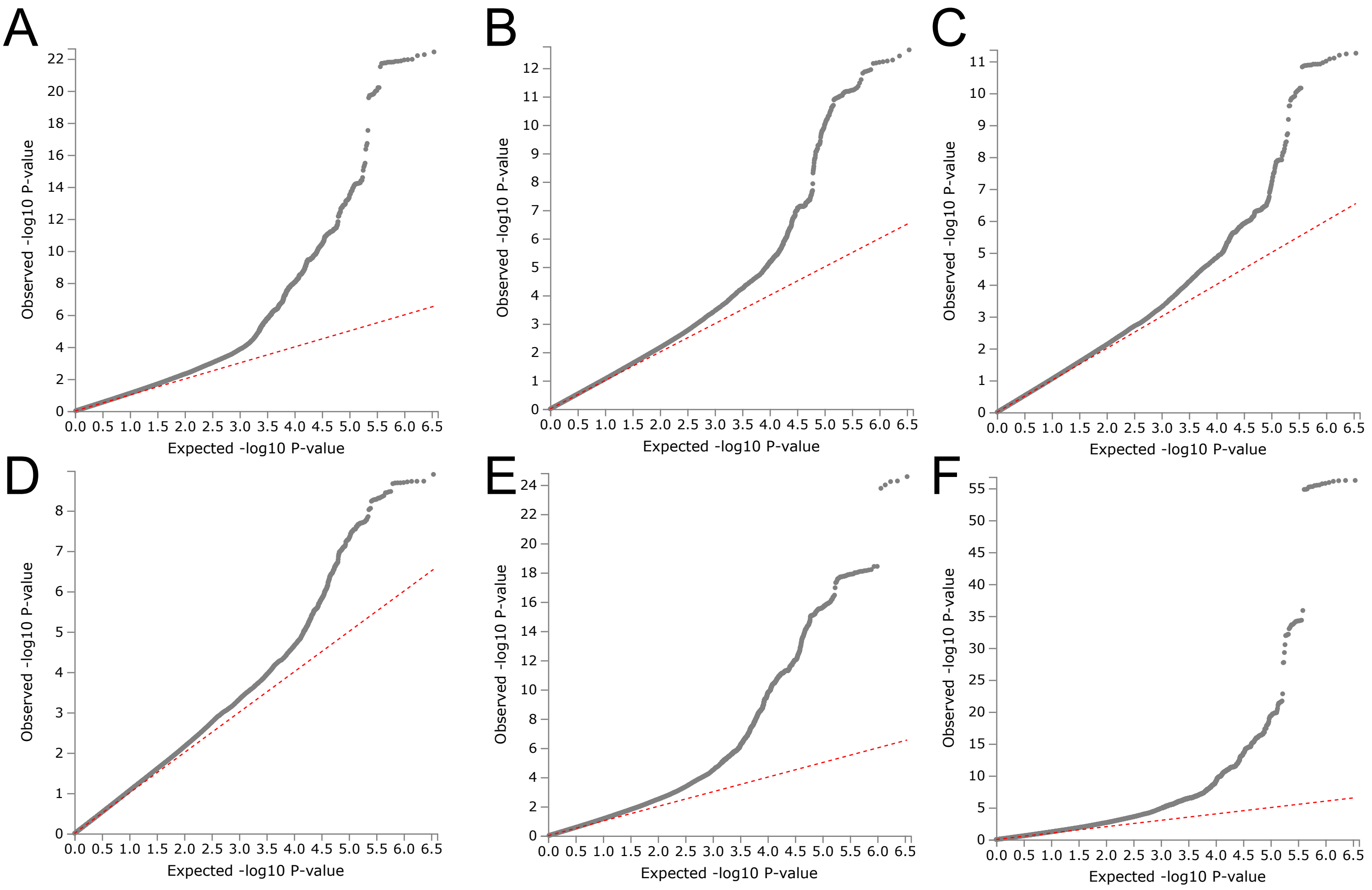


Supplementary Figure 1. QQplot of PE GWAS for sex combined sample (A) female offspring (B), male offspring (C), maternal sample (D), PW GWAS (E), and BW GWAS (F)

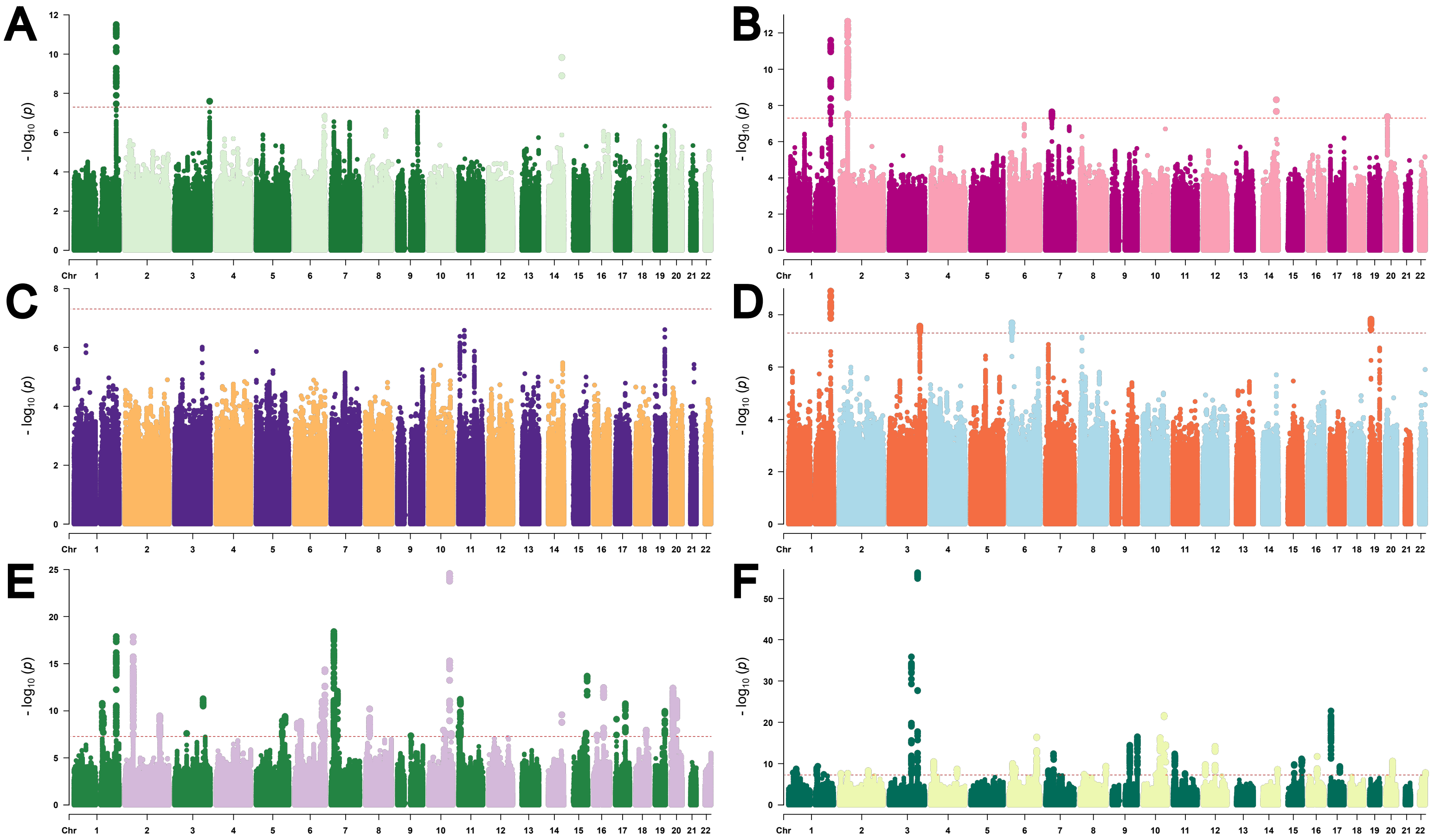

**Supplementary Figure 2.** Manhattan plot with the significant SNPs for male offspring (A), female offspring (B), paternal (C) and maternal (D) PLE GWAS, and PW (E) and BW GWAS (F). Each dot represents a genetic variant, plotted by chromosomal position (x-axis) and $-\log_{10}(p)$ (y-axis), with higher values indicating greater statistical significance. The horizontal dashed red line denotes the genome-wide significance threshold (p = 5×10-8).

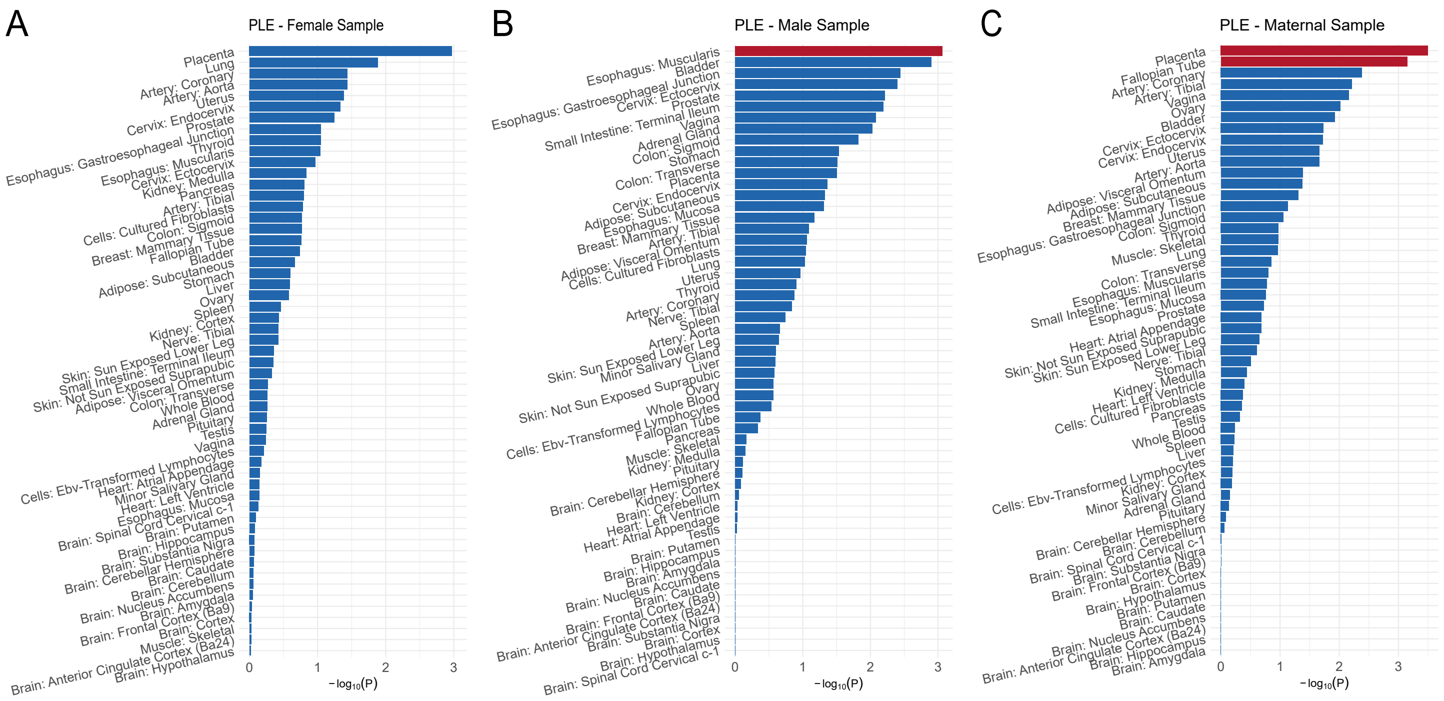


**Supplementary Figure 3: MAGMA tissue enrichment analysis for sex stratified and maternal GWAS sample.** MAGMA gene property analysis was performed on (A) Female PLE-GWAS, (B) Male PLE-GWAS and (C) Maternal-GWAS. Significant expression (red bars) was discovered for the Male PLE-GWAS in the esophagus muscularis, and for the Maternal PLE-GWAS in the placenta and fallopian tubes. No significant tissue enrichment was discovered for the Female PLE-GWAS.

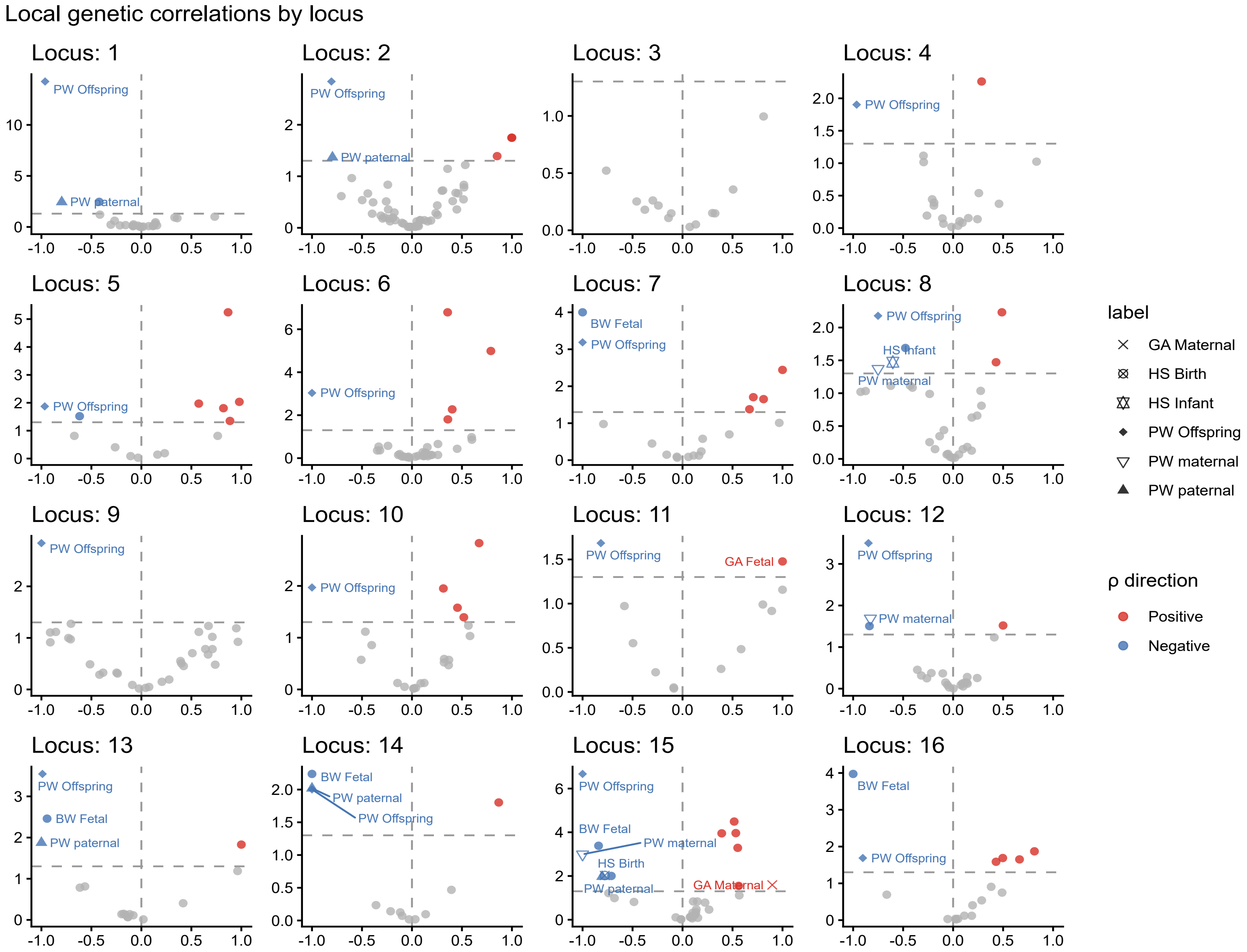


**Supplementary Figure 4.** Volcano plots of local genetic correlations in all mapped PLE, computed using LAVA. The x-axis shows local rg, and the y-axis shows -log10(P) values. Significant negative correlations are shown in blue and significant positive correlations are shown in red. Gray dashed lines indicate rg = 0 and the nominal significance threshold of p < .05, FDR corrected using Benjamini-Hochberg.

### Overlap of Significant Genes

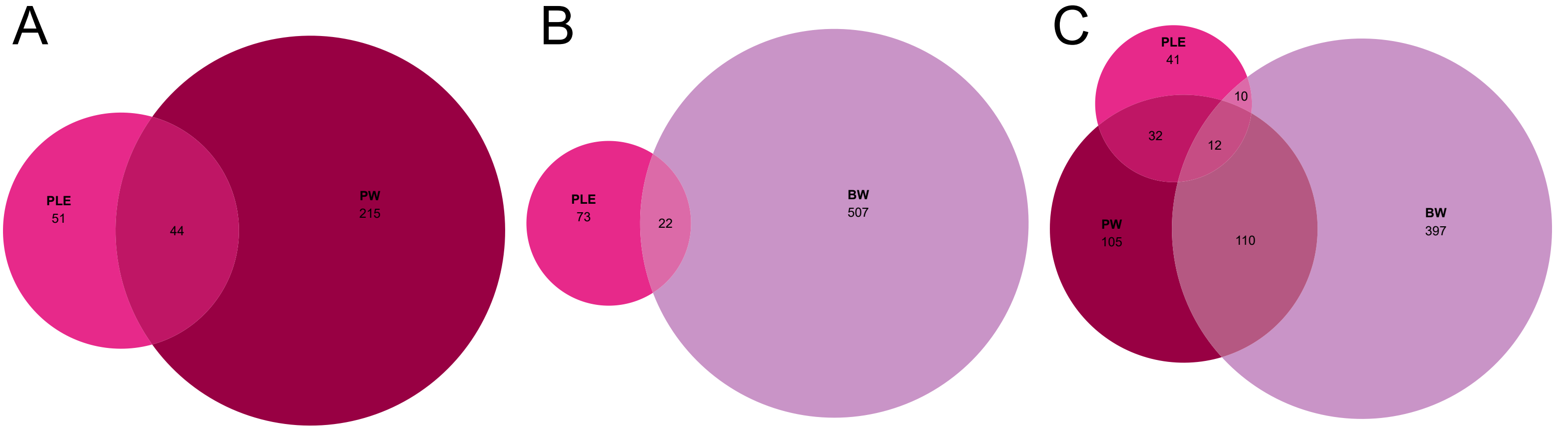


**Supplementary figure 5.** Overlap of significant genes between PLE and PW (A), PLE and BW (A), and PLE, PW, and BW (C).
