## Supplemental Results for "Genetic Architecture of Placental Efficiency for Term Infants: Monoaminergic Pathways and Placental Tissue Expression"

**Supplementary Results**

**Comparison of loci and genes identified in PLE, PW and BW GWAS**

To further characterize shared architecture, we compared lead SNPs and genomic risk loci across PLE, PW and BW offspring GWAS using Jaccard Index (JI) and Szymkiewicz–Simpson overlap coefficients (OC). On a phenotypic level, we observed the following correlations: BW and PW (r = 0.607), BW and PLE (r = 0.003), and PW and PLE (r = −0.76). Both JI and OC quantify similarity between two sets on a scale from 0-1, where higher values indicate stronger similarity. We observed no shared genome-wide significant SNPs between PLE and BW (JI = 0, OC = 0), whereas PLE and PW shared six lead SNPs (JI = 0.10, OC = 0.30). At the locus level, we observed a JI = 0.387, indicating that 38.7% of all base pairs were shared between PLE and PW, and an overlap coefficient of 0.709, indicating that 70.9% of PLE locus base pairs were contained within PW loci. In contrast, the JI for PLE and BW was 0.038, meaning that only 3.8% of base pairs overlapped, with an overlap coefficient of 0.102, showing that 10.2% of PLE base pairs were contained within BW loci. Locus-level comparisons further showed that 87.5% of PLE loci contained PW SNPs, while 58.3% of PW loci contained PLE SNPs, corresponding to an average locus-level overlap of 72.9%.

To assess overlap at the gene level we applied FDR correction to raw MAGMA p-values and assessed the overlap. At the gene level, we compared FDR-significant genes identified through MAGMA analysis, which revealed 44 overlapping genes between PLE (n = 51 unique genes) and PW (n = 215 unique genes; See **Supplementary Figure 4**). Taken together, these results suggest that while PLE and PW share substantial global genetic architecture, the primary signals driving each trait, particularly the lead SNPs, remain largely distinct.
